# Biometry and volumetry in multi-centric fetal brain MRI: assessing the bias of super-resolution reconstruction

**DOI:** 10.1101/2024.09.23.24313965

**Authors:** Thomas Sanchez, Angeline Mihailov, Mériam Koob, Nadine Girard, Aurélie Manchon, Ignacio Valenzuela, Marta Gómez-Chiari, Gerard Martí Juan, Alexandre Pron, Elisenda Eixarch, Gemma Piella, Miguel A. González Ballester, Oscar Camara, Vincent Dunet, Guillaume Auzias, Meritxell Bach Cuadra

## Abstract

**Background:** Super-resolution reconstruction (SRR) of fetal brain magnetic resonance imaging has the potential to enable the development of new imaging biomarkers to better study *in utero* neurodevelopment. However, potential biases in 2D biometric and 3D volumetric measurements due to different SRR techniques remain understudied.

**Purpose:** To assess the consistency of biometric and volumetric measurements across three hospitals using three widely used SRR pipelines.

**Materials and Methods:** This retrospective study used T2-weighted (T2w) fetal brain MRI scans acquired in routine clinical practice at three hospitals. MRIs from each subject were reconstructed with each of the 3 SRR methods. Four experts did biometric measurements on each SRR volume blinded to the method used. Automated 3D volumetry was performed using a state-of-the-art segmentation method. A univariate analysis was first carried out with Friedman tests with post-hoc Wilcoxon rank-sum tests, and results were confirmed in a multivariate analysis accounting for the effect of gestational age and different raters, using a t-distributed generalized additive model. An additional qualitative evaluation was performed to assess how likely clinicians would be to use the current SRR volumes in their practice, and whether they would prefer it to low-resolution T2w acquisitions. Differences were assessed with Friedman tests and post-hoc Wilcoxon rank-sum tests.

**Results:** 84 healthy subjects were included in three gestational age groups ([21-28): 25.4±1.9, [28-32): 29.3±1.3, [32-36): 33.5±1.2). Statistically significant differences in biometric measurements were found, but consistently remained below voxel width (0.8 mm). Automated 3D volumetry revealed systematic but very small effects (<2.8%). The qualitative evaluation showed systematic differences between SRR methods for the perception of white matter intensity (p=0.02) and sharpness of the image (p=0.01).

**Conclusion:** Variations in 2D and 3D quantitative measurements did not show any large systematic bias when using different SRR methods for radiological assessment in clinical routine across multiple centers, scanners, and raters.

**Summary:** Different super-resolution reconstruction methods for fetal brain MRI volumes lead to negligible variations in 2D or 3D quantitative measurements; this may help achieve larger sample sizes in prenatal development studies.

**Key Results:**

- In this multi-centric retrospective study, 252 super-resolution reconstructions (SRR) scans from 84 healthy subjects showed negligible variations in 2D in biometric measures (below the voxel with of 0.8 mm; p<0.001).
- 3D measurements revealed small variations ranging from 0.8 % in supratentorial tissues (p<0.001) to 2.8% in the extra-cerebral cerebrospinal fluid (p<0.001).
- Clinicians favored having both low resolution and SRR volumes available.

## Introduction

Fetal brain Magnetic Resonance Imaging (MRI) is increasingly used as a complement to ultrasound (US) imaging for confirming or ruling out equivocal findings^1^. Its excellent soft tissue contrast and image resolution enables more accurate measurements of the fetal brain as well as a better parenchymal signal, critical for detecting cortical malformations and subtle white matter anomalies^2^.

Antenatal brain MRI routine assessment combines qualitative morphological evaluation and biometric measurements. In routine clinical practice, fetal brain MRI biometry is performed on T2-weighted (T2w) stacks of two-dimensional slices with 2-5 mm thickness and 0.5-1 mm in-plane resolution, usually acquired following three orthogonal planes. However, fetal and maternal motion can lead to oblique acquisition planes, which, combined with the anisotropic image resolution, can make it difficult to carry out precise biometric measurements. Although some studies have compared measurements done on MRI to US reference values ^3–7^ used to establish deviation from normality, MRI-based biometric measurements are still not recommended in clinical practice because of the challenge of acquiring a precise slice orientation with MRI.

In the past decade, super-resolution reconstruction (SRR) methods^8–14^ have emerged, allowing the combination of motion-corrupted, low-resolution (LR) T2w series into a high-resolution 3D isotropic volume. These 3D volumes are valuable for fetal brain biometry, since they enable flexible navigation in any plane, facilitating the selection of optimal planes for precise biometric measurements^15–17^. Moreover, they enable a volumetric (3D) analysis, supported by several automated pipelines^8,10,12–14,18^. These techniques pave the road towards a more accurate characterization of normal and pathological fetal neurodevelopment using MRI.

Early work on SRR 3D volumes have compared the consistency of their biometric measurements with those from US and LR slices ^16,19–21^. Kyriakopoulou et al.^16^ used SRR volumes reconstructed using the Slice-to-Volume Reconstruction method^8,10^ to build normative models of both biometric and volumetric structures. Khawam et al.^19^ studied the inter-rater reliability between biometric measurements on T2w series and MIALSRTK-reconstructed volumes^12,18^, while Lamon et al.^20^ focused on corpus callosum biometry, comparing US, T2w, and SRR volumes reconstructed using MIALSTRK^12,18^. However, these works relied on a single SRR method, thus its replication with other SRR methods remains to be proven. Recently, Ciceri et al.^21^ compared for the first time 2D biometry across multiple SRR methods (MIALSRTK^12,18^, NiftyMIC^13^, and SVRTK^10,22,23^), focusing on the 20-21 gestational weeks period. They showed that MIALSRTK and NiftyMIC achieved a good reconstruction success rate and were consistent with T2w series measurements, while SVRTK showed many failed reconstructions and was excluded.

However, these works were all limited to mono-centric data, and did not consider whether SRR methods could improve inter-rater reliability or if they introduced systematic biases in quantitative measurements. Ciceri et al^21^. did not disentangle the effect of data quality from the impact of the SRR algorithm. By conflating the success rate of the compared SRR methods and the quality of the biometric measurements they could not answer the following question: when different SRR methods yield good quality results, will the biometric measurement values remain consistent? Or, framed differently: does the reconstruction process of any SRR method introduce alterations that systematically bias the biometric evaluation, even when the SRR is of good quality?

We hypothesized that given high-quality reconstructions, 2D and 3D measurements would be consistent across different SRR methods, but that experts would remain cautious about using SRR reconstructions for clinical assessments, because of alterations in the intensity of the reconstructed image. The purpose of this study was to evaluate the clinical usefulness of SRR and assess whether these methods could introduce artifacts that would systematically bias measurements taken from the reconstructed volumes.

## Materials and methods

### Dataset

#### Population

Brain MRI examinations were retrospectively collected from ongoing research studies at the three hospitals: Hospital Clínic de Barcelona (Barcelona, Spain), La Timone (Marseilles, France) and Lausanne University Hospital (CHUV, Lausanne, Switzerland). Exclusion criteria included twin pregnancies and any pathology or malformation in the fetal MRI scans. The study received ethical approval from each center’s institutional review board (CHUV: CER-VD 2021-00124, La Timone: Aix-Marseille University N°2022-04-14-003, Hospital Clínic: HCB/2022/0533). Fetal examinations were equally distributed across three gestational age (GA) bins representing different stages of fetal brain development: [21, 28) weeks, [28, 32) weeks and [32, 36) weeks. A flow diagram of included and excluded MRI examinations is shown in Figure 1.a.

**Figure 1.**
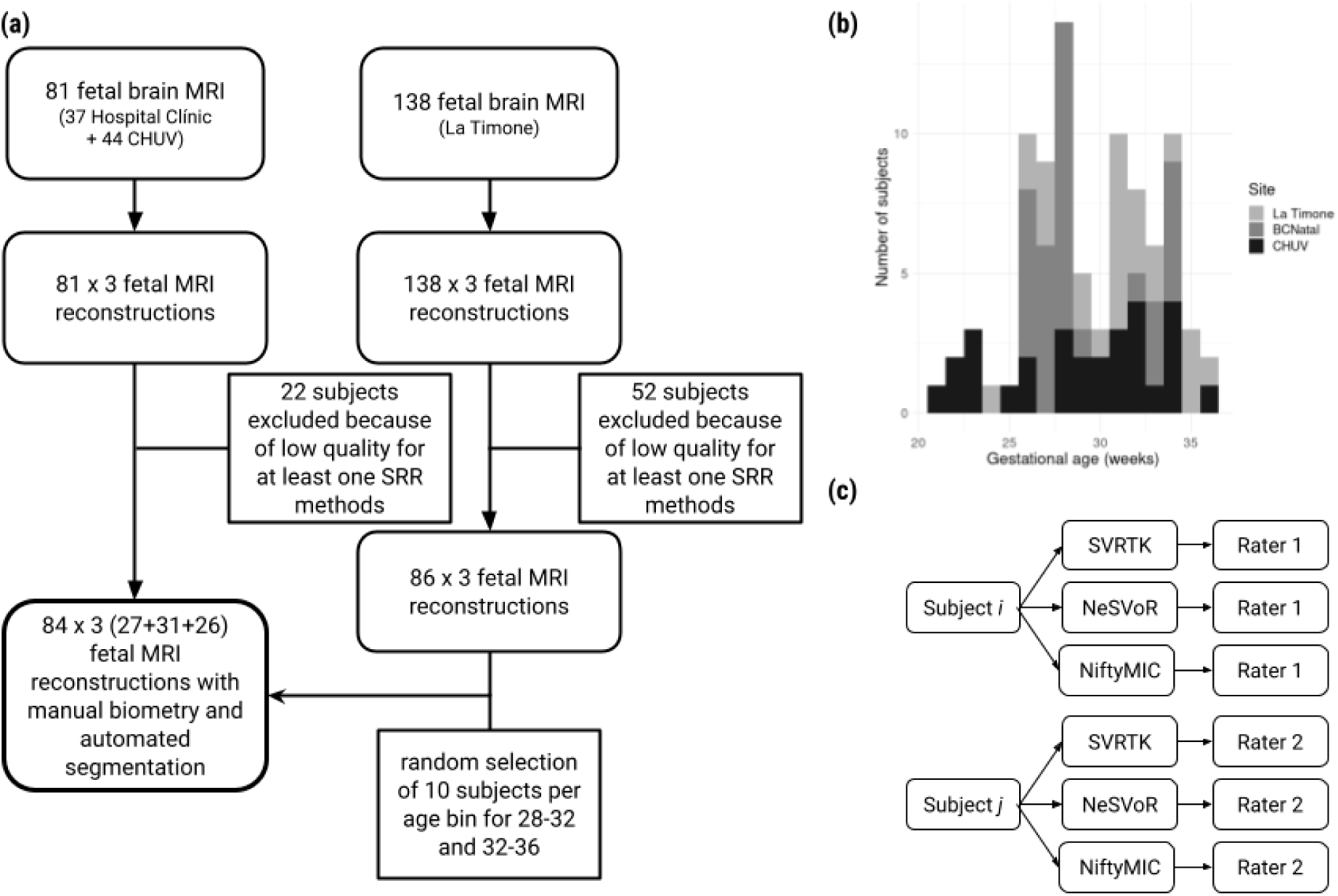
**(a)** Flowchart of our study sample shows inclusion and exclusion. There was a total of 219 pregnant patients who were imaged across three centers. Seventy-four MRI examinations were excluded due to poor-quality reconstruction, resulting in 145 MRI examinations that were annotated and automatically segmented. After selection of subjects in relevant age bins, this resulted in 84 MRI examinations analyzed (27 for ages [21-28) 31 for [28,32) and 26 for [32-36)). **(b)** Distribution of gestational ages across the different sites. **(c)** Design of the study. The subjects are nested within the raters. The raters considered the subjects from their center (NG, AM for La Timone, IV for Hospital Clínic, MK for CHUV) and performed the measurements on every reconstruction for each subject.

#### MRI Data

Fetal MRI data were acquired with different Siemens scanners (Erlangen, Germany) at 1.5T or 3T across hospitals. The fetal brain MRI protocol included T2w HASTE (Half-Fourier Acquisition Single-shot Turbo spin Echo imaging) sequences acquired in three orthogonal directions (axial, coronal, sagittal). Details on the different MRI acquisition parameters, and number of acquisitions per subject are available in Table 1.

**Table 1.** Metadata regarding the acquisition parameters, the gestational ages of participants, the resolution of the T2w series and the number stacks used in the reconstruction algorithm.

| Site | Scanner | Field [T] | $n_{\text{sub}}$ | 21-28 | 28-32 | 32-36 | LR resolution [mm<sup>3</sup>] | $n_{\text{stacks}}$ |
| --- | --- | --- | --- | --- | --- | --- | --- | --- |
| CHUV | Aera | 1.5 | 19 | 9 | 8 | 2 | 1.12 x 1.12 x 3.3 | 6.2±3.1 |
|  | MAGNETOM Sola | 1.5 | 8 | 0 | 3 | 5 | 1.12 x 1.12 x 3.3 | 6.3±1.3 |
|  | Skyra | 3 | 2 | 0 | 0 | 2 | 0.55 x 0.55 x 3.0 | 5.5±2.1 |
| Hospital Clínic | Aera | 1.5 | 29 | 12 | 10 | 7 | 0.55 x 0.55 x 2.8 | 12.0±3.3 |
| La Timone | SymphonyTim | 1.5 | 17 | 3 | 6 | 8 | 0.74 x 0.74 x 3.5 | 4.7± 1.8 |
|  | Skyra | 3 | 9 | 3 | 4 | 2 | 0.68 x 0.68 x 3.0 | 3.4±0.7 |

#### MRI data processing

As clinical fetal brain MRI acquisitions feature anisotropic resolution, the data acquired in different orientations are reconstructed into a single, high-resolution volume through SRR methods. Each subject was reconstructed using three widely used SRR toolkits: NeSVoR (v.0.5.0)^14^, NiftyMIC (v.0.9.0)^13^, and SVRTK (v.auto-2.2.0)^10,22,23^. Depending on the hospital, stacks with high levels of motion or signal drops were excluded through visual inspection^19^ and/or automated quality control^24^. At La Timone and Hospital Clínic, stacks were processed with non-local means denoising^25^ and N4 bias field correction^26^. Each subject was then reconstructed using the default parameters of the three SRR methods, at 0.8mm isotropic resolution. The resulting SRR volumes were aligned to a standard orientation.

For poor quality reconstructions, different stacks combinations were tested until the image quality was deemed sufficient by visual assessment (no evident artifacts or errors from registration/reconstruction). If no combination resulted in a sufficiently high-quality reconstruction, the subject was excluded from the study.

### Biometric Measurements

Biometric measurements were performed on both LR 2D stacks and 3D SRR volumes using ITK-SNAP (University of Pennsylvania, PA, USA). Measures were performed on each site by medical experts in obstetric and/or pediatric image analysis: IV (5 years of experience) for Hospital Clínic, NG (> 20 years of experience) and AM (5 years of experience) for La Timone and MK (15 years of experience) for CHUV. This resulted in a design where subjects are nested within the raters (Fig. 1.c.). Following established guidelines for fetal brain MRI biometry^1,3,16,27^, the following measurements were performed: length of the corpus callosum (LCC), height of the vermis (HV), brain and skull biparietal diameters (bBIP, sBIP), and transverse cerebellar diameter (TCD). An example of the measurements on a subject is shown in Figure 2. These measurements were then compared to the reference values obtained by Kyriakopoulou et al.^16^

**Figure 2.**
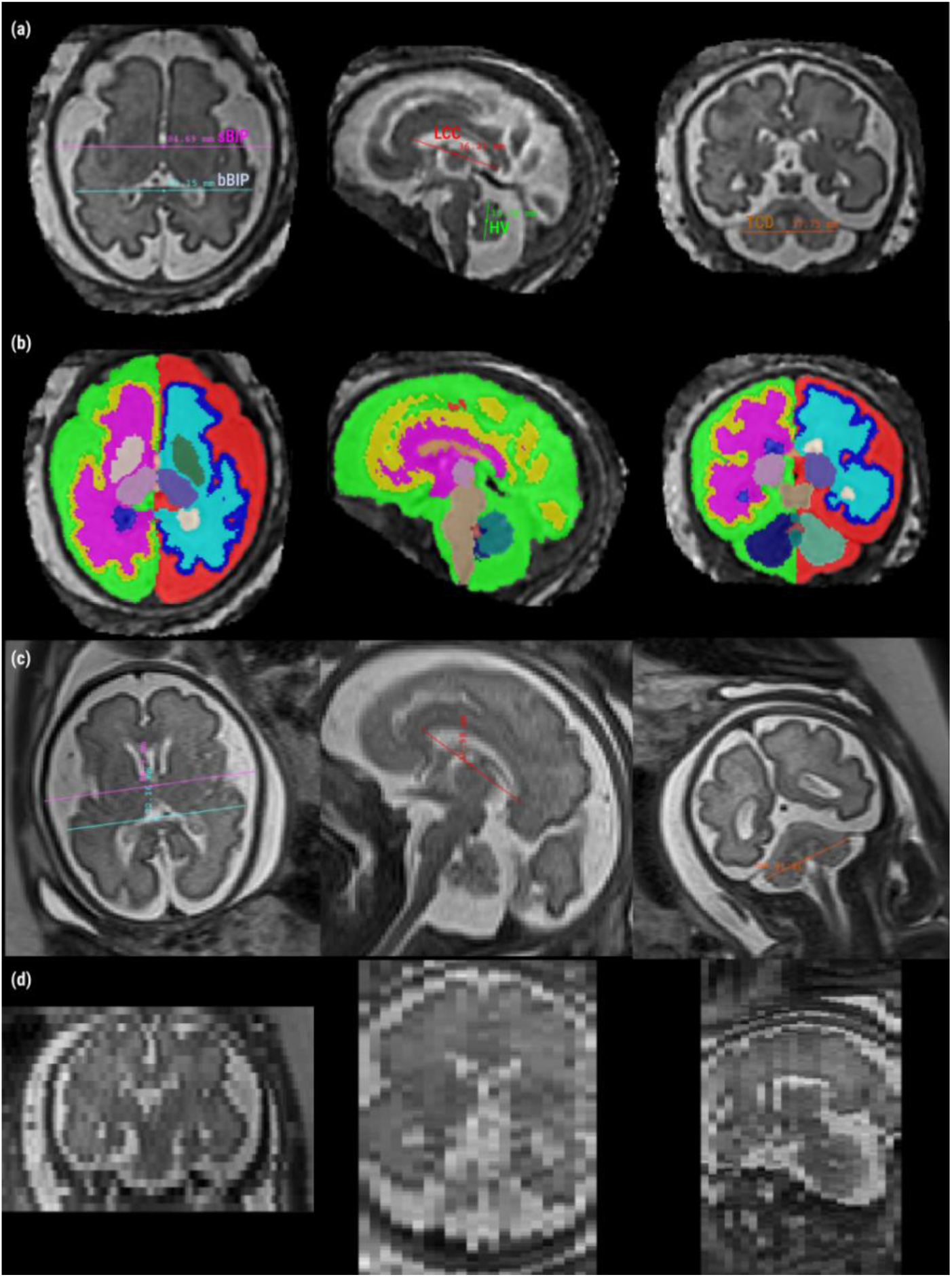
2D measurements guidelines. **(a)** Measurements done on a 31-week-old subject, reconstructed using SVRTK. Axial: brain and skull biparietal diameters (bBIP and sBIP). Sagittal: length of the corpus callosum (LCC) and height of the vermis (HV). Coronal: transverse cerebellar diameter (TCD). **(b)** Automated segmentation using BOUNTI **(c)** Measurements on the T2w stacks. Each column represents a different stack. The stacks were re-oriented for visualization purposes **(d)** Through-plane view of the low-resolution images of (c), showing the thick slices of the LR acquisitions.

On the LR stacks, each rater chose the stack best suited (in terms of alignment and image quality) for each measurement. On the 3D SRR volumes, raters had the option to re-align (manual rigid transformation) the images prior to performing the measurements. In total, the four different raters each performed around 550 measurements (5 structures x 4 variants (1 LR + 3 SRR) x 26-29 subjects).

#### Automated volumetry

Automated volumetric evaluation was carried out on the SRR reconstructed volumes using BOUNTI^28^, a recent deep learning segmentation method. BOUNTI segments the brain into 19 different regions and was trained on a large corpus of manually segmented brains volumes. An illustration of the segmentations is provided in Figure 2b. In our analysis, we considered five volumetric measurements for which reference values are available^16^: extra-cerebral cerebrospinal fluid (eCSF), cortical gray matter (cGM), cerebellum (CBM), supratentorial brain tissue (ST) and total lateral ventricles (VT). cGM and CBM measurements were also compared to the growth curves from Machado-Rivas et al.^29^, which used the methods of Kainz et al.^11^ to reconstruct the T2w stacks, and automated segmentation with an atlas-based approach^15^.

#### Qualitative assessment

We aimed at obtaining expert feedback on the appearance, particularly on the aspects of intensity and visibility, of key anatomical structures used to assess fetal development. Four neuroradiologists (NG, >20 years of experience; AM, 5 years of experience; MG,12 years of experience; MK, 15 years of experience, were asked to qualitatively assess the volumes reconstructed from six subjects using all three SRR methods considered. The subjects were selected to represent different GA bins (26, 28, 29, 30, 32, and 34 weeks) with high quality 3D SRR volumes for all subjects and methods to avoid any bias. In a first round of evaluation, the clinicians visualized all SRR volumes from a given subject and were asked to assess how clearly different structures appeared in the SRR volume. The details of the questions asked, and structures rated are available in supplementary Table S9. In a second stage, raters were asked to compare the SRR volumes from each subject with the corresponding LR stacks of images. They were first asked to rank the three SRR volumes for each subject based on their likelihood of use (with ties allowed). They were then asked to determine whether they would choose the SRR volume over the LR stacks for their clinical assessment, and whether the SRR volume provided more information than the LR stacks for a radiological evaluation.

### Statistical analysis

A univariate analysis was initially carried out to assess the influence of the SRR algorithm on the biometric (respectively volumetric) measurements. Due to the non-Gaussian distribution of the data, a Friedman test (the non-parametric equivalent of a repeated measures ANOVA, N=252, degrees of freedom=2) was used to test the difference across SRR methods. We did not apply corrections for multiple comparisons to detect even small statistical effects related to the SRR techniques, as correction would actually make it easier to support our hypothesis. Post-hoc testing was done using pairwise Wilcoxon rank-sum tests, and Bonferroni correction for multiple comparisons was applied at this stage. Effect sizes were reported as 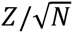.

We confirmed these results using multivariate regression to evaluate the impact of SRR on biometric (resp. volumetric) measurements while accounting for covariates. A t-distributed Generalized Additive Model for Scale and Location (GAMLSS)^30,31^ was fitted with the biometric (resp. volumetric) measurement as the response, the SRR algorithm as the fixed effect of interest, gestational age (GA) as a covariate, rater as a covariate for the biometry only (as the volumetry is computed automatically), and subject as a random effect.

The choice of a GAMLSS model over a simpler t-distributed linear mixed effect (LME) model was based on visual inspection of the residual distribution (R function fitdistrplus::descdist) and of the cumulative distribution function (R function DHARMa::simulateResiduals). While both the LME and the GAMLSS had a well-aligned cumulative distribution function, the GAMLSS model showed a less dispersed residual distribution, suggesting more stable estimates.

The qualitative analysis relied on a smaller sample. We nonetheless carried out a univariate analysis using a Friedman test (N=72, degrees of freedom=2). When significant results were found, post-hoc analysis testing was done using pairwise Wilcoxon rank-sum tests, with Bonferroni correction for multiple comparisons. All statistical analyses were carried out using the R software (version 4.2.2). To facilitate the analysis of the results, the ratings of AM were used in a confirmatory analysis as part of a supplementary experiment. The analysis then simply has subjects nested within raters.

## Results

### Population

After application of the inclusion and exclusion criteria (Figure 1.a.), 252 SRR from 84 healthy fetuses were included: 29 at the Hospital Clínic, 26 at La Timone and 29 at CHUV. The distribution of gestational age is shown in Figure 1.b. and broken down by age bins in Table 1.

### Biometry measurements across SR reconstruction methods

Univariate and multivariate statistics are reported in Table 2. There was no significant difference induced by SRR methods on LCC and HV in the univariate analysis, very small effects in the multivariate analysis, –0.2±0.06 mm (p < 0.001) for the NeSVoR-NiftyMIC difference in LCC, −0.09±0.94 (p < 0.05) for the NeSVoR-SVRTK difference in HV. When comparisons yielded statistically significant results, the effect sizes systematically remained small (at most 0.43±0.06 mm for the sBIP), smaller than a 0.1% variation and below the width of a voxel (0.8mm).

**Table 2.**
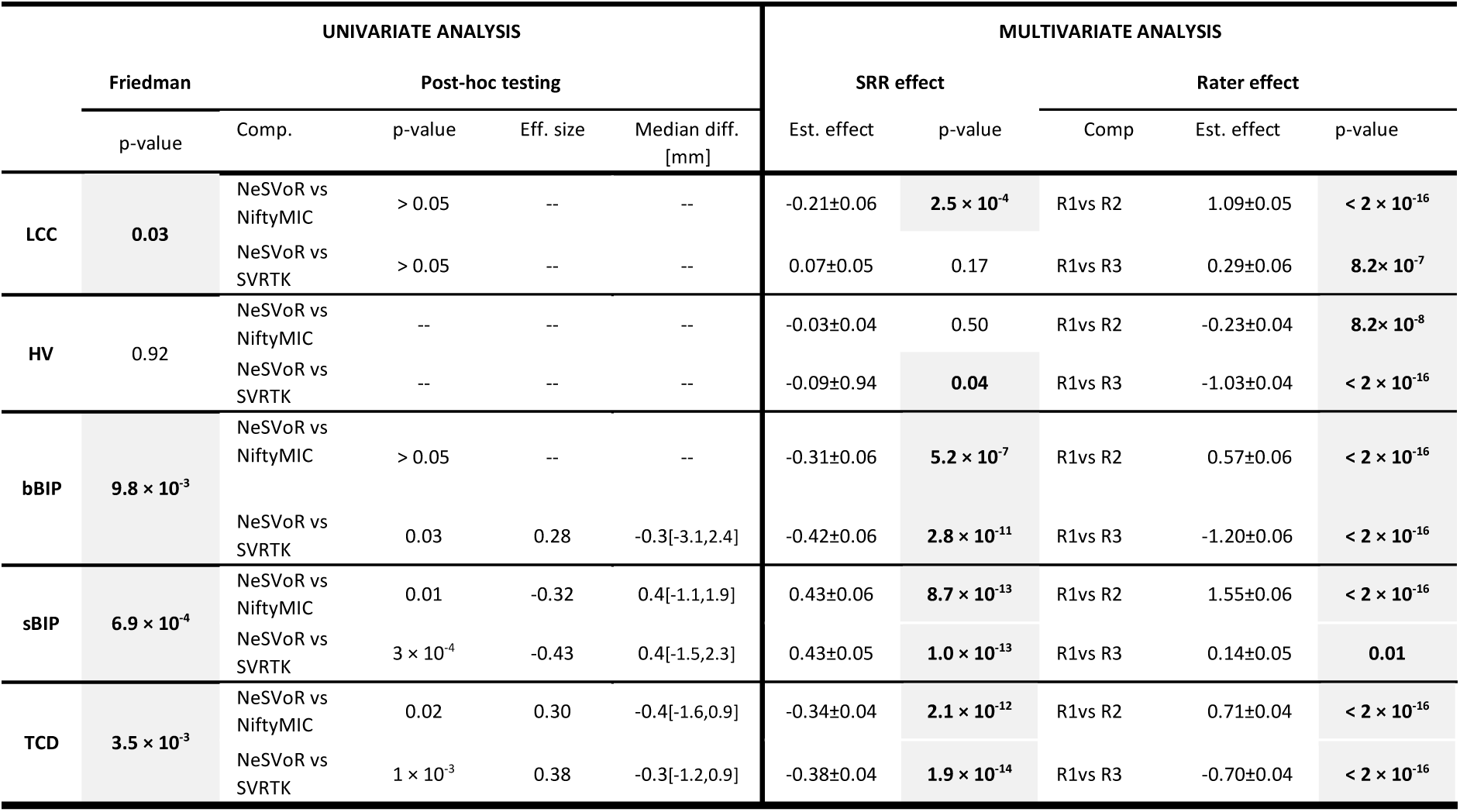
Statistical analyses for biometry measurements. Univariate biometry analysis (N= 252, df =2) and multivariate biometry analysis using a t-distributed GAMLSS model.

| UNIVARIATE ANALYSIS |  |  |  |  |  | MULTIVARIATE ANALYSIS |  |  |  |  |
| --- | --- | --- | --- | --- | --- | --- | --- | --- | --- | --- |
|  | Friedman | Post-hoc testing |  |  |  | SRR effect |  | Rater effect |  |  |
|  | p-value | Comp. | p-value | Eff. size | Median diff. [mm] | Est. effect | p-value | Comp. | Est. effect | p-value |
| LCC | 0.03 | NeSVoR vs NiftyMIC | > 0.05 | -- | -- | -0.21±0.06 | $2.5 \times 10^{-4}$ | R1vs R2 | 1.09±0.05 | $< 2 \times 10^{-16}$ |
| | | NeSVoR vs SVRTK | > 0.05 | -- | -- | 0.07±0.05 | 0.17 | R1vs R3 | 0.29±0.06 | $8.2 \times 10^{-7}$ |
| HV | 0.92 | NeSVoR vs NiftyMIC | -- | -- | -- | -0.03±0.04 | 0.50 | R1vs R2 | -0.23±0.04 | $8.2 \times 10^{-8}$ |
| | | NeSVoR vs SVRTK | -- | -- | -- | -0.09±0.94 | 0.04 | R1vs R3 | -1.03±0.04 | $< 2 \times 10^{-16}$ |
| bBIP | $9.8 \times 10^{-3}$ | NeSVoR vs NiftyMIC | > 0.05 | -- | -- | -0.31±0.06 | $5.2 \times 10^{-7}$ | R1vs R2 | 0.57±0.06 | $< 2 \times 10^{-16}$ |
| | | NeSVoR vs SVRTK | 0.03 | 0.28 | -0.3[-3.1,2.4] | -0.42±0.06 | $2.8 \times 10^{-11}$ | R1vs R3 | -1.20±0.06 | $< 2 \times 10^{-16}$ |
| sBIP | $6.9 \times 10^{-4}$ | NeSVoR vs NiftyMIC | 0.01 | -0.32 | 0.4[-1.1,1.9] | 0.43±0.06 | $8.7 \times 10^{-13}$ | R1vs R2 | 1.55±0.06 | $< 2 \times 10^{-16}$ |
| | | NeSVoR vs SVRTK | $3 \times 10^{-4}$ | -0.43 | 0.4[-1.5,2.3] | 0.43±0.05 | $1.0 \times 10^{-13}$ | R1vs R3 | 0.14±0.05 | 0.01 |
| TCD | $3.5 \times 10^{-3}$ | NeSVoR vs NiftyMIC | 0.02 | 0.30 | -0.4[-1.6,0.9] | -0.34±0.04 | $2.1 \times 10^{-12}$ | R1vs R2 | 0.71±0.04 | $< 2 \times 10^{-16}$ |
| | | NeSVoR vs SVRTK | $1 \times 10^{-3}$ | 0.38 | -0.3[-1.2,0.9] | -0.38±0.04 | $1.9 \times 10^{-14}$ | R1vs R3 | -0.70±0.04 | $< 2 \times 10^{-16}$ |

The multivariate analysis also allowed estimating effects related to the raters, which were consistently larger than the SRR effects, but remained small. The effect was at most 1.55 mm for sBIP (2.5% variability). These results were confirmed by an additional, single-site analysis, where two raters annotated the same data (see Supplementary materials).

Growth charts are provided in Figure 3 (top row) and in line with the centiles estimated in previous works^3,16,32^. Further illustration of the different growth curves for the different raters and SRR are provided in Supplementary Figure S1.

**Figure 3.**
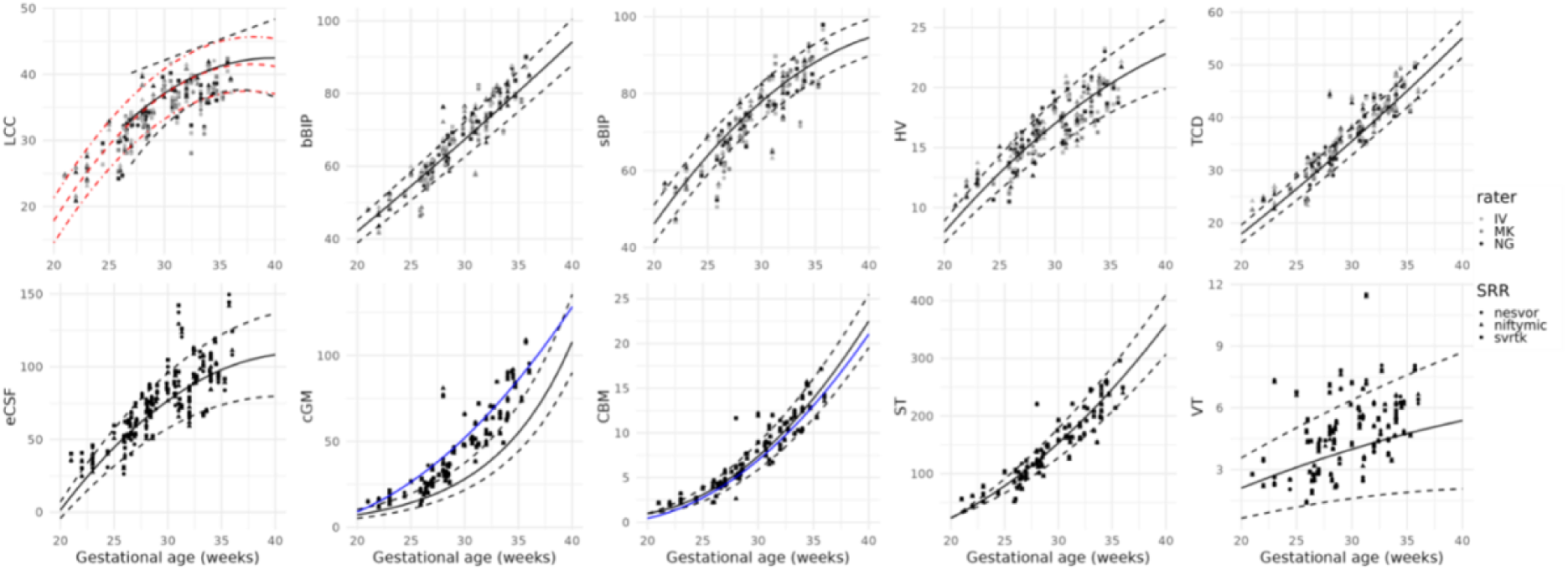
Top row. Biometric measurements as a function of gestational age, for the different SRR methods and raters. The curves and dashed lines represent normative 5^th^, 50^th^ and 95^th^ centiles from Kyriakopoulou et al.^16^, except for LCC, where the black curve is from measurements on HASTE acquisitions from Tilea et al. (2009)^3^ and the red one from ultrasound measurements done by Pashaj et al. (2013)^31^. **Bottom row.** Volumetric measures as a function of gestational age, for the different SRR methods and sites. The curves and dashed lines represent normative 5^th^, 50^th^ and 95^th^ centiles from Kyriakopoulou et al.^27^ and additional blue curves are taken from Machado-Rivas et al.^28^.

#### Brain tissue volumetry

Results for automated brain tissue volumetry are provided in Table 3 and show a small but consistent variability between SRR methods, in the order of 1%, except for eCSF, where 2.7% differences were observed between NeSVoR and NiftyMIC.

**Table 3.** Statistical analyses for volumetry measurements. Univariate biometry analysis (N= 252, df =2) and multivariate biometry analysis using a t-distributed GAMLSS model.

|  | UNIVARIATE ANALYSIS |  |  |  |  | MULTIVARIATE ANALYSIS |  |  |
| --- | --- | --- | --- | --- | --- | --- | --- | --- |
|  | Friedman |  | Post-hoc testing |  |  | SRR effect |  |  |
|  | p-value | Comparison | p-value | Effect | Median diff. [cm'] | Comparison | Est. effect | p-value |
| eCSF | 5.5×10<sup>-11</sup> | NeSVoR vs NiftyMIC | >0.05 | -- |  | NeSVoR vs NiftyMIC | -1.84±0.16 | < 2 × 10<sup>-16</sup> |
|  |  | NeSVoR vs SVRTK | 1×10<sup>-4</sup> | 0.45 | -1.8[-12.8,2.3] | NeSVoR vs SVRTK | -0.19±0.18 | 0.31 |
|  |  | NiftyMIC vs SVRTK | 7×10<sup>-13</sup> | 0.80 | 2.1[-0.4,10.8] |  |  |  |
| cGM | 4.9×10<sup>-14</sup> | NeSVoR vs NiftyMIC | 3×10<sup>-9</sup> | 0.67 | 0.7[-0.7,2.6] | NeSVoR vs NiftyMIC | -0.68±0.03 | < 2 × 10<sup>-16</sup> |
|  |  | NeSVoR vs SVRTK | 7×10<sup>-8</sup> | 0.61 | 0.5[-0.7,2.2] | NeSVoR vs SVRTK | -0.39±0.03 | < 2 × 10<sup>-16</sup> |
|  |  | NiftyMIC vs SVRTK | 0.003 | 0.36 | 0.3[-1.5,1.4] |  |  |  |
| CBM | 7.5 × 10<sup>-6</sup> | NeSVoR vs NiftyMIC | > 0.05 | -- |  | NeSVoR vs NiftyMIC | -0.04±0.01 | 1 × 10<sup>-13</sup> |
|  |  | NeSVoR vs SVRTK | 3×10<sup>-4</sup> | 0.42 | 0.1[-0.2, 0.3] | NeSVoR vs SVRTK | -0.02±0.01 | 0.001 |
|  |  | NiftyMIC vs SVRTK | 2×10<sup>-5</sup> | 0.49 | 0.05[-0.1,0.3] |  |  |  |
| ST | 6.1 × 10<sup>-12</sup> | NeSVoR vs NiftyMIC | 3×10<sup>-10</sup> | 0.71 | 1.2[-0.7, 4.7] | NeSVoR vs NiftyMIC | -0.84±0.07 | < 2 × 10<sup>-16</sup> |
|  |  | NeSVoR vs SVRTK | 0.03 | 0.29 | 0.3[-1.5,1.8] | NeSVoR vs SVRTK | -0.43±0.06 | 7 × 10<sup>-12</sup> |
|  |  | NiftyMIC vs SVRTK | 1×10<sup>-6</sup> | 0.55 | 0.5[-0.7,4.0] |  |  |  |
| VT | 1.9 × 10<sup>-7</sup> | NeSVoR vs NiftyMIC | 7×10<sup>-6</sup> | 0.52 | 0.1[-0.2, 0.3] | NeSVoR vs NiftyMIC | -0.06±0.01 | < 2 × 10<sup>-16</sup> |
|  |  | NeSVoR vs SVRTK | 0.005 | 0.34 | 0.1[-0.1, 0.3] | NeSVoR vs SVRTK | -0.03±0.01 | 9 × 10<sup>-7</sup> |
|  |  | NiftyMIC vs SVRTK | 0.02 | 0.39 | 0.1[-0.2, 0.1] |  |  |  |

Growth curves for volumetry are provided in Figure 3 (bottom row) and yield values that generally align with previously estimated centiles^16^, except for the cortical gray matter, which was consistently overestimated compared to Kyriakopoulou et al.^16^, and underestimated compared to Machado-Rivas et al.^29^.

#### Qualitative feedback on SRR

In the first qualitative experiment evaluating the presence and visibility of specific anatomical structures on SRR volumes, clinicians rated most volumes from NeSVoR and NiftyMIC as insufficient for their radiological assessment. While SVRTK images were rated of sufficiently good quality (better quality than NeSVoR, p=0.013), clinicians remained hesitant to use them in a radiological assessment. An excerpt from the results is shown in Table 4A, where we see that while all SRR methods yield good cortical continuity and sharpness, NeSVoR performed poorly on the white matter (layering: SVRTK-NeSVoR=0.5 (p=0.004), intensity: SVRTK-NeSVoR=0.63 (p=0.01), NiftyMIC-NeSVoR = 0.54 (p=0.003)) and is blurrier than SVRTK and NiftyMIC (blurriness: SVRTK-NeSVoR=0.84 (p=0.001), NiftyMIC – NeSVoR =0.62 (p=0.02)), leading to an overall worse perceived quality (quality: SVRTK-NeSVoR=0.63 (p=0.01)). Additional results on the corpus callosum, ventricles, internal capsule and posterior fossa are available in the supplementary material. An example of reconstructions is shown in Figure 4, where the worst and best rated SRR volumes are presented side-by-side, along with the acquired LR stacks in the three orientations. Overall, NeSVoR was often graded lower than SVRTK and NiftyMIC due to alterations introduced by the method in the white matter homogeneity and intensity (Figure 4, subject 1). On the other hand, the best rated volume (Figure 4, subject 2 with SVRTK) has a very clear white matter, with a marked contrast between the white matter and the basal ganglia.

**Figure 4.**
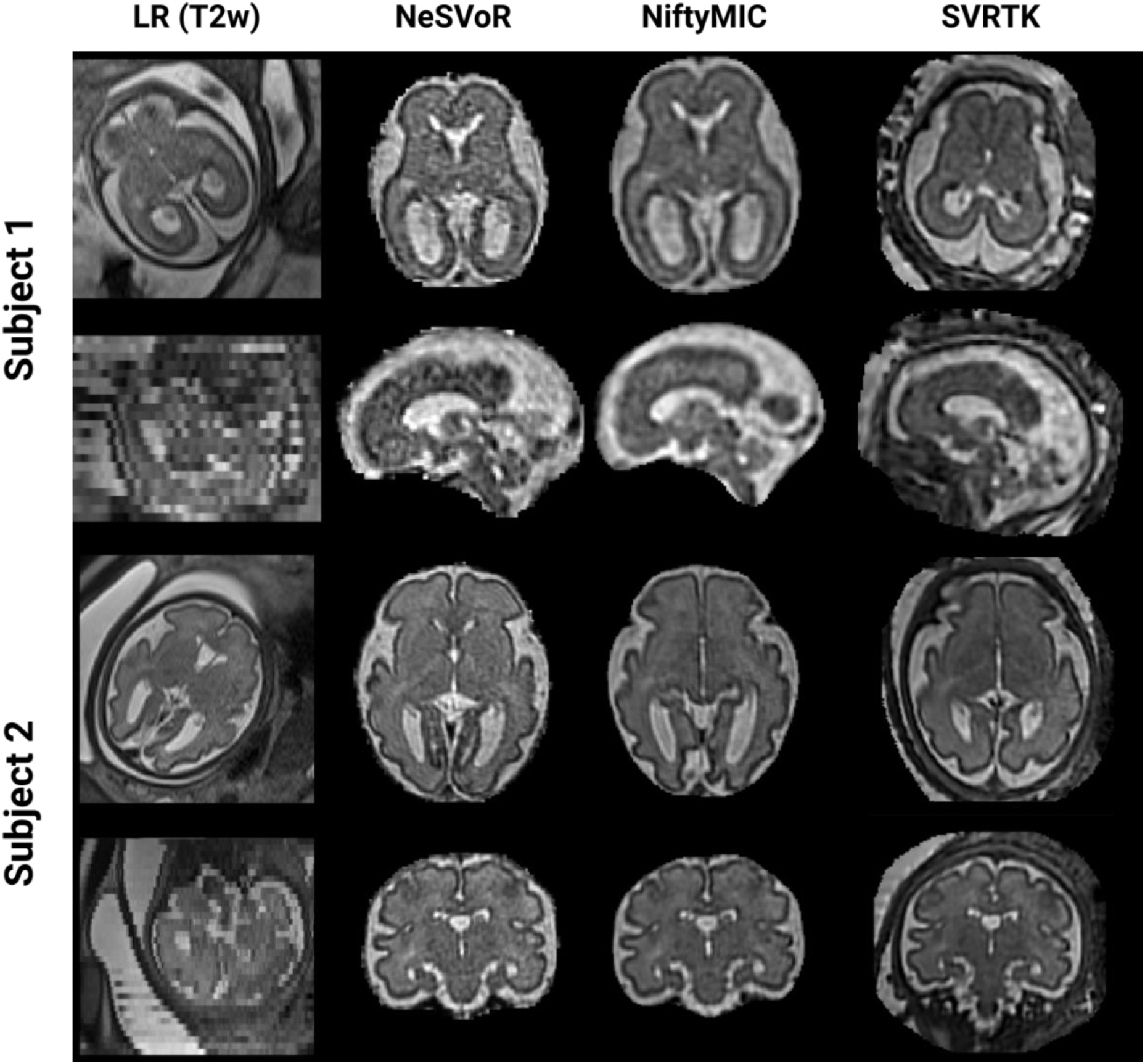
Example of two subjects (GA=26w and 30w) with in-plane views of three different T2w acquisitions along with the reconstructed volumes. On the top, subject 1 reconstructed with NeSVoR is the worst rated SR volume (global subjective quality = 0) and on the bottom, subject 2 reconstructed with SVRTK is the best rated SR volume (global subjective quality = 1.54).

**Table 4.** Top. Subjective structural quality assessment. Scores range between 0 (bad), 1 (acceptable) and 2 (excellent). A single star means that the method is statistically significantly better than the worst performing method of the column. **Bottom.** Qualitative comparison between SR and LR. Scores range from 0 to 2, the first column reflects a ranking, the second refer to whether the clinician would use SRR instead of LR volumes (choose only one), and the last column refer to whether the SRR was judged more suited for their clinical examination than LR. A score of 1 means that SRR is as useful as LR.

| (A) | Cortex |  | White matter |  | Global |  |
| --- | --- | --- | --- | --- | --- | --- |
|  | Continuity | Sharpness | Layering | Intensity | Blurriness | Quality |
| NeSVoR | 1.50±0.54 | 1.50±0.52 | 0.83±0.56 | 0.54±0.36 | 0.58±0.43 | 0.54±0.58 |
| NiftyMIC | 1.58±0.50 | 1.54±0.30 | 1.17±0.64 | 1.08±0.59* | 1.20±0.52* | 0.88±0.70 |
| SVRTK | 1.50±0.46 | 1.65±0.37 | 1.33±0.37* | 1.17±0.49* | 1.42±0.39* | 1.17±0.50* |
| (B) | SRR ranking |  | SRR *instead* of LR? |  | SRR *better* than LR? |  |
| NeSVoR | 0.58±0.52 |  | 0.79±0.52 |  | 0.79±0.72 |  |
| NiftyMIC | 1.46±0.72* |  | 1.21±0.72 |  | 1.17±0.63 |  |
| SVRTK | 1.17±0.71 |  | 1.08±0.76 |  | 1.08±0.78 |  |

In the second experiment (Table 4B), the raters ranked the different SRR volumes between each other, and the LR stacks. The results showed that the NeSVoR reconstructions were consistently rated lower than NiftyMIC and SVRTK, with NiftyMIC rated best in this experiment (SRR ranking: NiftyMIC-NeSVoR=0.86 (p=0.004)). When compared to the LR stacks, there was no unanimous preference for SRR volumes over LR images. Experts noted that most of the NiftyMIC and SVRTK volumes were considered usable as LR images but were rather hesitant in using NeSVoR instead of the LR images for their evaluation.

## Discussion

Today, advanced image processing techniques such as motion estimation and SRR allow us to freely navigate in 3D into the fetal brain to extract quantitative measurements. The aim of our study was to assess whether different state-of-the-art SRR methods induced systematic biases when reconstructed volumes are used for biometric and volumetric analyses. Results from multi-centric, multi-scanner acquisitions show statistically significant differences in 2D biometry across SRR methods, with differences consistently remaining below the voxel width (0.8 mm). On 3D volumetric measurements, trends are similar, with deviations in the order of 1% (2.5% for eCSF, due to different ways of cropping the brain across SRR methods). While small, the deviations in volumetry are systematic and might be a concern for future fine-grained analyses. Larger deviations from reference growth curves were observed for the cortical gray matter, where even results from Kyriakopoulou et al.^16^ and Machado-Rivas et al.^29^ exhibited large variations. This is likely due to differences in reconstruction and segmentation protocols between these two works as well as the data used to train the BOUNTI model^28^, as variations in the manual delineation of cGM are notoriously hard to control^33^.

Our work supplements the study of Ciceri et al.^21^, who showed in a more restricted setting (20-21 weeks, mono-centric) the consistency of the measurements done on two SRR methods. Our results are reassuring towards using SRR volumes in clinical practice or leveraging and comparing results from different studies: even if different SRR methods were to be deployed in clinical practice or used in multi-centric studies, biometric and volumetric measurements would remain consistent across sites, thus opening the door to new biomarkers, which cannot be obtained from US or LR stacks.

In addition, while SRR could be readily used for quantitative measurements, challenges remain due to the differences introduced by SRR methods (textured noise, intensity variations), which can appear depending on the original resolution settings. In our experiments, this is particularly pronounced in the case of NeSVoR. Therefore, training physicians to distinguish between SR reconstruction artifacts and structural alterations would be paramount when making SRR widely available. Nevertheless, clinicians generally agreed on the benefits of having *both* LR and SRR volumes available. This could help in detecting cortical malformations, as the gyrification is more clearly visible on SRR data since navigating in 3D in SRR data helps to reduce ambiguities caused by the uncontrolled sampling with 2D slices with LR stacks.

This work also shows that the true benefits of SRR would be revealed for biometric measurements of structure that require a precise anatomical orientation. This is the case for median structures like the length of the corpus callosum or the height of the vermis.

Nevertheless, despite this multi-centric and multi-rater study, our work should be further extended to include a holistic evaluation of the reconstructed volumes, notably including their quality and their ability to reconstruct pathological subjects. This would be necessary to truly assess the potential of these reconstruction methods in clinical settings. Overall, our study indicates that, when comparable 3D SR volumes of sufficient quality are achieved, the choice of SRR method does not introduce large systematic biases in 2D or 3D measurements.

## Data Availability

All data produced in the present study are available upon reasonable request to the authors

## Acknowledgements

This work was funded by Era-net NEURON MULTIFACT project (TS: Swiss National Science Foundation grant 31NE30_203977; AM, GA: French National Research Agency, Grant ANR-21-NEU2-0005; IV, EE: Instituto de Salud Carlos III (ISCIII) grant AC21_2/00016, GM, MG, OC, GP: Ministry of Science, Innovation and Universities: MCIN/AEI/10.13039/501100011033/), and the SulcalGRIDS Project, (GA: French National Research Agency Grant ANR-19-CE45-0014).

## Supplementary material

### Intra-rater reliability between LR and SRR biometry measurements

#### Materials and methods

Intra-rater reliability was evaluated using Lin’s Concordance Correlation Coefficient^34^.

#### Results

In Table S1, intra-rater reliability is reported for the three raters considered. CCC is very high for most structures (above 0.9) indicating very strong reliability. The lowest scores (although still high) are obtained for median structures (length of corpus callosum and height of the vermis). There is no major concern that a given SRR method would lead to a decrease in agreement between the SRR and LR. Figure S1 provides a visual comparison with the Pearson correlation coefficient and shows clearly that LCC and HV have more scattered measures compared to bBIP, sBIP and TCD. Moreover, some bias in the measurements can be observed from IV and MK in the LCC, and NG in the HV. This is not surprising given that obtaining precise planes for measurements is challenging in LR stacks.

**Table S1.** Lin’s Concordance Correlation Coefficient (CCC) between the LR and SR measurements for each rater. This supplements the results presented in Figure 3. Measurements with CCC below 0.9 are highlighted in blue.

|  | IV |  |  | MK |  |  | NG |  |  |
| --- | --- | --- | --- | --- | --- | --- | --- | --- | --- |
|  | NeSVoR | NiftyMIC | SVRTK | NeSVoR | NiftyMIC | SVRTK | NeSVoR | NiftyMIC | SVRTK |
| LCC | 0.73 | 0.65 | 0.69 | 0.87 | 0.86 | 0.86 | 0.93 | 0.92 | 0.92 |
| HV | 0.91 | 0.91 | 0.90 | 0.92 | 0.92 | 0.93 | 0.87 | 0.89 | 0.90 |
| bBIP | 0.99 | 0.99 | 0.99 | 0.99 | 0.99 | 0.98 | 0.98 | 0.98 | 0.98 |
| sBIP | 0.98 | 0.99 | 0.98 | 0.99 | 0.99 | 0.99 | 0.99 | 0.99 | 0.99 |
| TCD | 0.98 | 0.98 | 0.97 | 0.99 | 0.99 | 0.99 | 0.97 | 0.98 | 0.98 |

**Figure S1.**
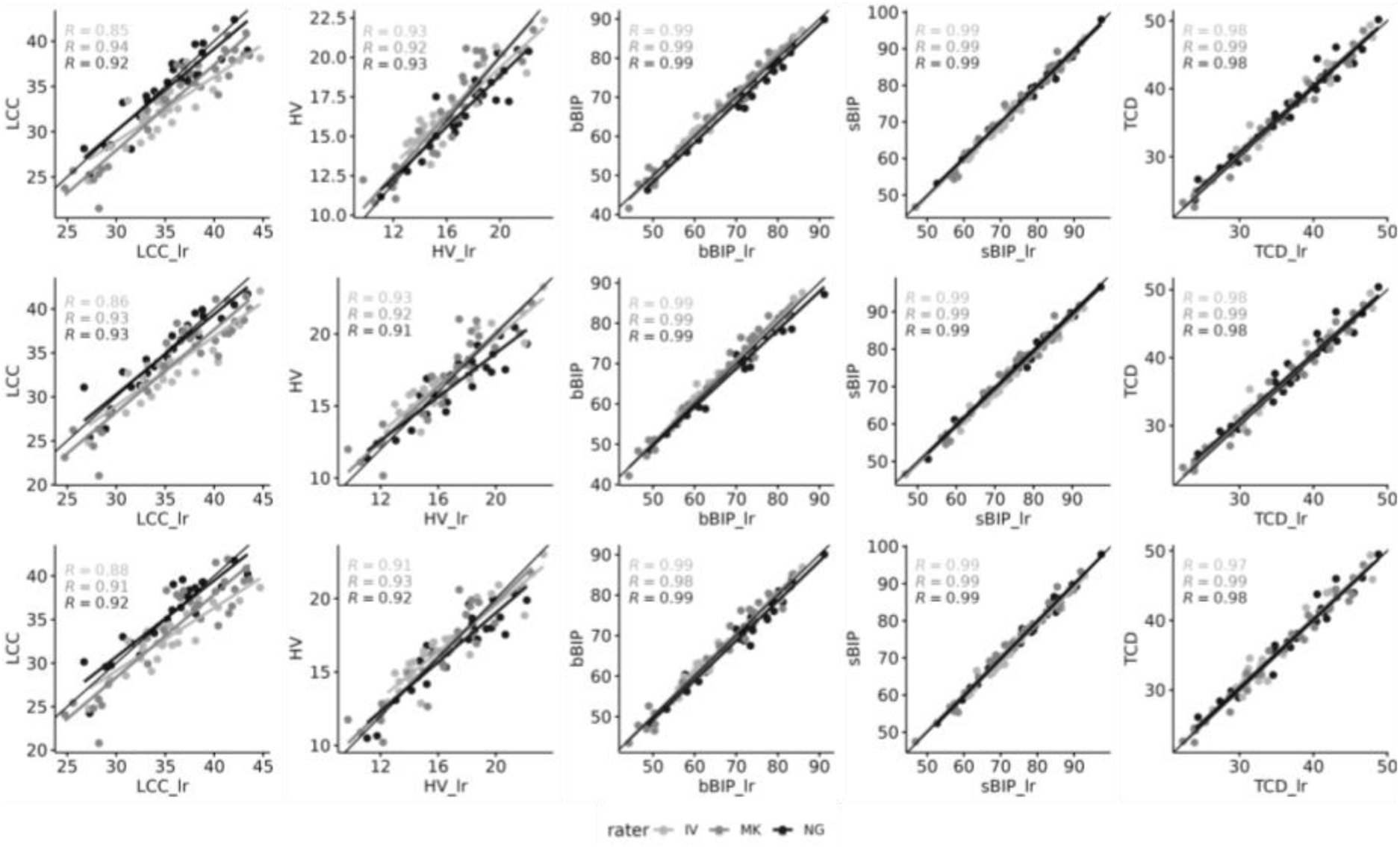
Linear regression between the LR and SR measurements for each rater.

### Complete statistical results for volumetry and biometry

Tables S2 and S3 contain the univariate and multivariate analyses for the biometry, and Tables S3 and S4 contain the univariate and multivariate analyses for the volumetry experiment.

**Table S2.** Statistical analyses for biometry measurements. Univariate analysis N= 252, df =2.

| | Friedman $\chi^2$ | p-value | Post-hoc testing | | | | |
| --- | --- | --- | --- | --- | --- | --- | --- |
|  |  |  | Comparison | p-value | Eff. size | Median diff. [mm] | Median abs. diff [mm] |
| **LCC** | 6.93 | 0.03 | Non-significant after correction for multiple testing |  |  |  |  |
| **HV** | 0.17 | 0.92 |  |  |  |  |  |
| **bBIP** | 9.24 | $9.8 \times 10^{-3}$ | NeSVoR vs SVRTK | 0.03 | 0.28 | 0.3[-2.4, 3.1] | 0.7[0.2,3.4] |
| **sBIP** | 14.55 | $6.9 \times 10^{-4}$ | NeSVoR vs SVRTK | $3 \times 10^{-4}$ | 0.43 | -0.4[-1.9, 1.1] | 0.7[0.1,2.4] |
|  |  |  | NeSVoR vs NiftyMIC | 0.01 | 0.32 | -0.4[-2.3,1.5] | 0.8[0.1,2.2] |
| **TCD** | 11.31 | $3.5 \times 10^{-3}$ | NeSVoR vs SVRTK | $1 \times 10^{-3}$ | 0.38 | 0.4[-0.9, 1.6] | 0.6[0.1,1.6] |
|  |  |  | NeSVoR vs NiftyMIC | 0.02 | 0.30 | 0.3[-0.9, 1.2] | 0.4[0.03,1.8] |

**Table S3.** Statistical analyses for biometry measurements. Multivariate analysis using a t-distributed GAMLSS model.

|  | Comparison | SRR effect |  |  | Comp. | Rater effect |  |  |
| --- | --- | --- | --- | --- | --- | --- | --- | --- |
|  |  | Est. effect [mm] | t-val. | p-value |  | Est. effect [mm] | t-val. | p-value |
| **LCC** | NeSVoR vs NiftyMIC | -0.31±0.10 | -3.04 | **0.003** | R1 vs R2 | 1.85±0.11 | -3.04 | **0.003** |
|  | NeSVoR vs SVRTK | -0.06±0.10 | 0.65 | 0.51 | R1 vs R3 | 0.54±0.11 | 5.06 | **1.1 × 10<sup>-6</sup>** |
| **HV** | NeSVoR vs NiftyMIC | -0.05±0.07 | -0.75 | 0.45 | R1 vs R2 | -0.17±0.07 | -2.43 | **0.01** |
|  | NeSVoR vs SVRTK | -0.08±0.07 | -1.28 | 0.20 | R1 vs R3 | -0.99±0.07 | -14.06 | **< 2 × 10<sup>-16</sup>** |
| **bBIP** | NeSVoR vs NiftyMIC | -0.25±0.14 | -1.74 | 0.08 | R1 vs R2 | 0.56±0.15 | 3.84 | **1.7 × 10<sup>-4</sup>** |
|  | NeSVoR vs SVRTK | -0.35±0.14 | -2.48 | **0.01** | R1 vs R3 | -1.15±0.15 | -7.75 | **9 × 10<sup>-13</sup>** |
| **sBIP** | NeSVoR vs NiftyMIC | 0.38±0.09 | 4.04 | **8.2 × 10<sup>-5</sup>** | R1 vs R2 | 1.82±0.09 | 19.1 | **< 2 × 10<sup>-16</sup>** |
|  | NeSVoR vs SVRTK | 0.43±0.09 | 4.63 | **7.3 × 10<sup>-6</sup>** | R1 vs R3 | 0.32±0.09 | 4.04 | **0.001** |
| **TCD** | NeSVoR vs NiftyMIC | -0.22±0.07 | -3.33 | **0.001** | R1 vs R2 | 0.63±0.07 | 9.23 | **< 2 × 10<sup>-16</sup>** |
|  | NeSVoR vs SVRTK | -0.35±0.07 | -5.29 | **3.9 × 10<sup>-7</sup>** | R1 vs R3 | -0.63±0.07 | -9.06 | **5 × 10<sup>-16</sup>** |

**Table S4.** Statistical analyses for volumetry measurements. Univariate analysis (N= 252, df =2)

| | Friedman $\chi^2$ | p-value | Post-hoc testing | | | |
| --- | --- | --- | --- | --- | --- | --- |
|  |  |  | Comparison | p-value | Eff. size | Median diff. [cm<sup>3</sup>] |
| **eCSF** | 47.21 | 5.5×10<sup>-11</sup> | NeSVoR vs SVRTK | 1×10<sup>-4</sup> | 0.45 | -1.82[-12.83,2.28] |
|  |  |  | NiftyMIC vs SVRTK | 7×10<sup>-13</sup> | 0.80 | 2.11[-0.35,10.75] |
| **cGM** | 61.31 | 4.9×10<sup>-14</sup> | NeSVoR vs NiftyMIC | 3×10<sup>-9</sup> | 0.67 | 0.66[-0.74,2.58] |
|  |  |  | NeSVoR vs SVRTK | 7×10<sup>-8</sup> | 0.61 | 0.46[-0.69,2.23] |
|  |  |  | NiftyMIC vs SVRTK | 0.003 | 0.36 | 0.30[-1.54,1.40] |
| **CBM** | 23.60 | 7.5 × 10<sup>-6</sup> | NeSVoR vs SVRTK | 3×10<sup>-4</sup> | 0.42 | 0.06[-0.16, 0.32] |
|  |  |  | NiftyMIC vs SVRTK | 2×10<sup>-5</sup> | 0.49 | 0.04[-0.10,0.33] |
| **ST** | 51.63 | 6.1 × 10<sup>-12</sup> | NeSVoR vs NiftyMIC | 3×10<sup>-10</sup> | 0.71 | 1.16[-0.69, 4.68] |
|  |  |  | NeSVoR vs SVRTK | 0.03 | 0.29 | 0.34[-1.45,1.84] |
|  |  |  | NiftyMIC vs SVRTK | 1×10<sup>-6</sup> | 0.55 | 0.48[-0.69,3.95] |
| **VT** | 30.93 | 1.9 × 10<sup>-7</sup> | NeSVoR vs NiftyMIC | 7×10<sup>-6</sup> | 0.52 | 0.07[-0.18, 0.27] |
|  |  |  | NeSVoR vs SVRTK | 0.005 | 0.34 | 0.05[-0.14, 0.25] |
|  |  |  | NiftyMIC vs SVRTK | 0.02 | 0.39 | 0.05[-0.22, 0.12] |

**Table S5.** Statistical analyses for volumetry measurements. Multivariate analysis using a t-distributed GAMLSS model.

| **SRR effect** |  |  |  |  |
| --- | --- | --- | --- | --- |
|  | Comparison | Est. Effect<br>[cm<sup>3</sup>] | t-val. | p-value |
| **eCSF** | NeSVoR vs NiftyMIC | -1.84±0.16 | -11.32 | **< 2 × 10<sup>-16</sup>** |
|  | NeSVoR vs SVRTK | -0.18±0.18 | -1.07 | 0.31 |
| **cGM** | NeSVoR vs NiftyMIC | -0.68±0.03 | -19.87 | **< 2 × 10<sup>-16</sup>** |
|  | NeSVoR vs SVRTK | -0.39±0.03 | -11.42 | **< 2 × 10<sup>-16</sup>** |
| **CBM** | NeSVoR vs NiftyMIC | -0.04±0.01 | -8.15 | **9 × 10<sup>-14</sup>** |
|  | NeSVoR vs SVRTK | -0.02±0.01 | -3.33 | **0.001** |
| **ST** | NeSVoR vs NiftyMIC | -0.84±0.07 | -11.88 | **< 2 × 10<sup>-16</sup>** |
|  | NeSVoR vs SVRTK | -0.43±0.06 | -7.40 | **8 × 10<sup>-12</sup>** |
| **VT** | NeSVoR vs NiftyMIC | -0.06±0.01 | -11.81 | **< 2 × 10<sup>-16</sup>** |
|  | NeSVoR vs SVRTK | -0.03±0.01 | -5.11 | **9 × 10<sup>-7</sup>** |

#### Single-site multi-rater analysis

As the data were rated twice at La Timone, this allowed us to carry out a more in-depth, single site analysis, removing potential confounders introduced by the nested design of the study. Tables S6, S7 and S8 respectively show the intra-and inter-rater reliability, the univariate biometric analysis and the multivariate analysis. The results are in line with the ones in the main paper, except that in this mono-centric evaluation, the effect of SRR is non-significant (the effect size remains the same).

The only additional result is the inter-rater reliability between AM and NG, which remains very high overall, although it is slightly lower on median structures, especially in LR vermis height.

**Table S6.** Intra and inter-rater reliability. Intra-rater reliability was evaluated using Lin’s Concordance Correlation Coefficient (CC) and inter-rater reliability was evaluated using two-way Intraclass Correlation Coefficient (ICC).

|  | Intra-rater reliability (LR-SRR) |  |  |  |  |  | Inter-rater reliability |  |  |  |
| --- | --- | --- | --- | --- | --- | --- | --- | --- | --- | --- |
|  | AM |  |  | NG |  |  | LR | NeSVoR | NiftyMIC | SVRTK |
|  | NeSVoR | NiftyMIC | SVRTK | NeSVoR | NiftyMIC | SVRTK |  |  |  |  |
| **LCC** | 0.95 | 0.93 | 0.95 | 0.93 | 0.92 | 0.92 | 0.96 | 0.96 | 0.97 | 0.93 |
| **HV** | 0.97 | 0.98 | 0.95 | 0.87 | 0.90 | 0.90 | 0.89 | 0.95 | 0.95 | 0.94 |
| **bBIP** | 0.99 | 0.99 | 0.99 | 0.98 | 0.98 | 0.98 | 0.99 | 0.99 | 0.99 | 0.99 |
| **sBIP** | 0.99 | 1.00 | 1.00 | 0.99 | 0.99 | 0.99 | 1.00 | 0.99 | 0.99 | 1.00 |
| **TCD** | 0.99 | 0.99 | 0.99 | 0.97 | 0.98 | 0.98 | 0.98 | 0.99 | 0.99 | 0.99 |

**Table S7.**
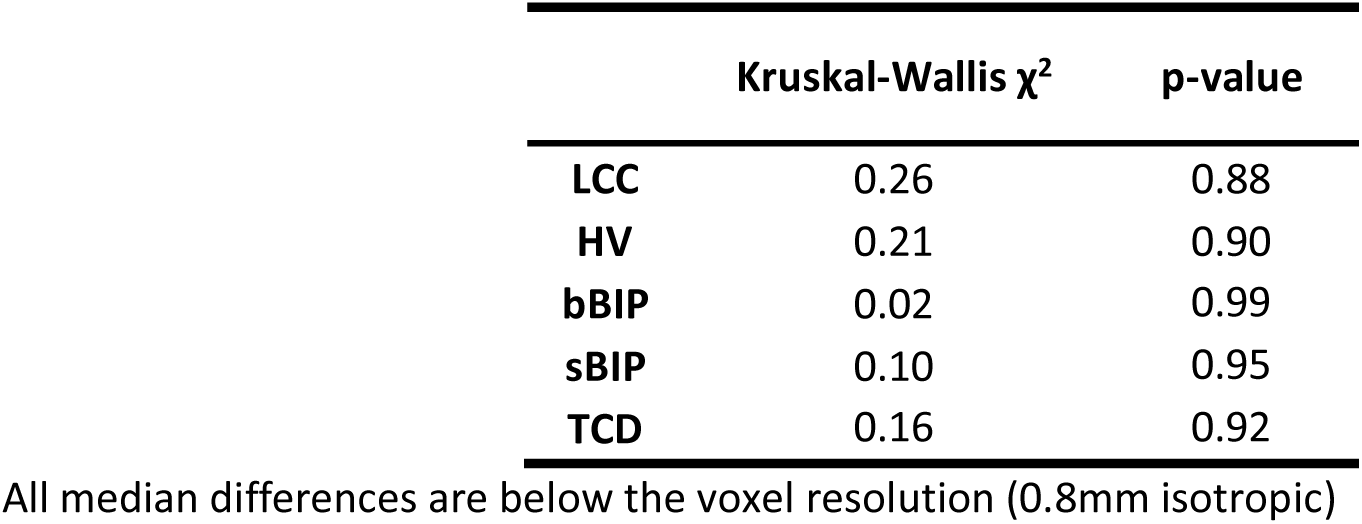
Univariate analysis – Single site and two raters - N=156, df =2. A Kruskal-Wallis test was chosen as Friedman test does not allow for replicated measurements.

| | Kruskal-Wallis $\chi^2$ | p-value |
| --- | --- | --- |
| **LCC** | 0.26 | 0.88 |
| **HV** | 0.21 | 0.90 |
| **bBIP** | 0.02 | 0.99 |
| **sBIP** | 0.10 | 0.95 |
| **TCD** | 0.16 | 0.92 |

All median differences are below the voxel resolution (0.8mm isotropic)

**Table S8.**
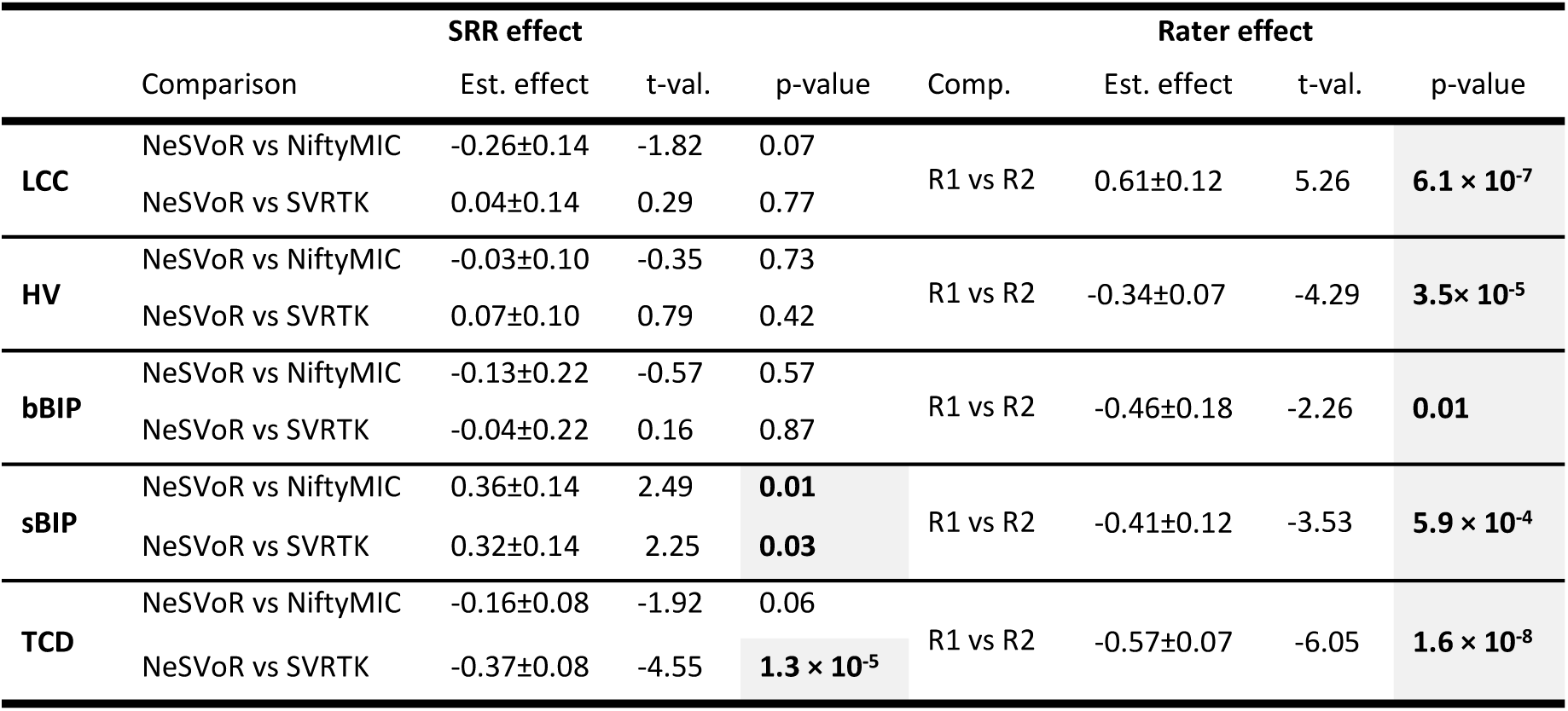
Multivariate analysis – Single site and two raters – t-distributed GAMLSS model.

### Rater-wise, SRR-wise regression predictions

In Figure S2, we present a visual representation of the fits obtained using the data from different raters and the different SRR methods. It shows visually how more variability in the prediction originates from the rater rather than the SRR method.

**Figure S2.**
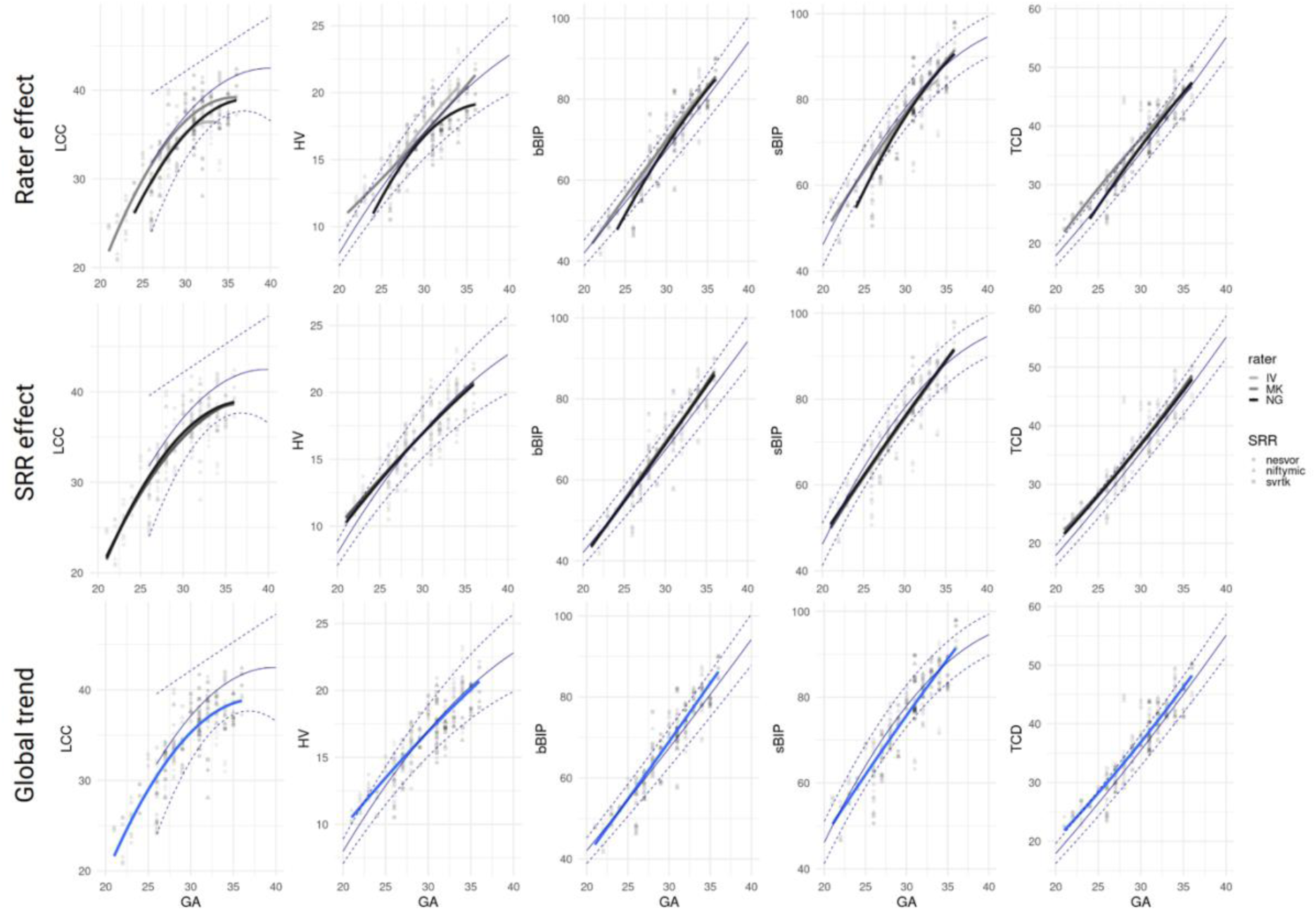
Quadratic fit split by rater (first row), by SRR method (second row) and global trend (third row). This visually illustrates the sources of variability in the fitting from different sources.

#### Additional results of the subjective rating experiment

**Table S9.**
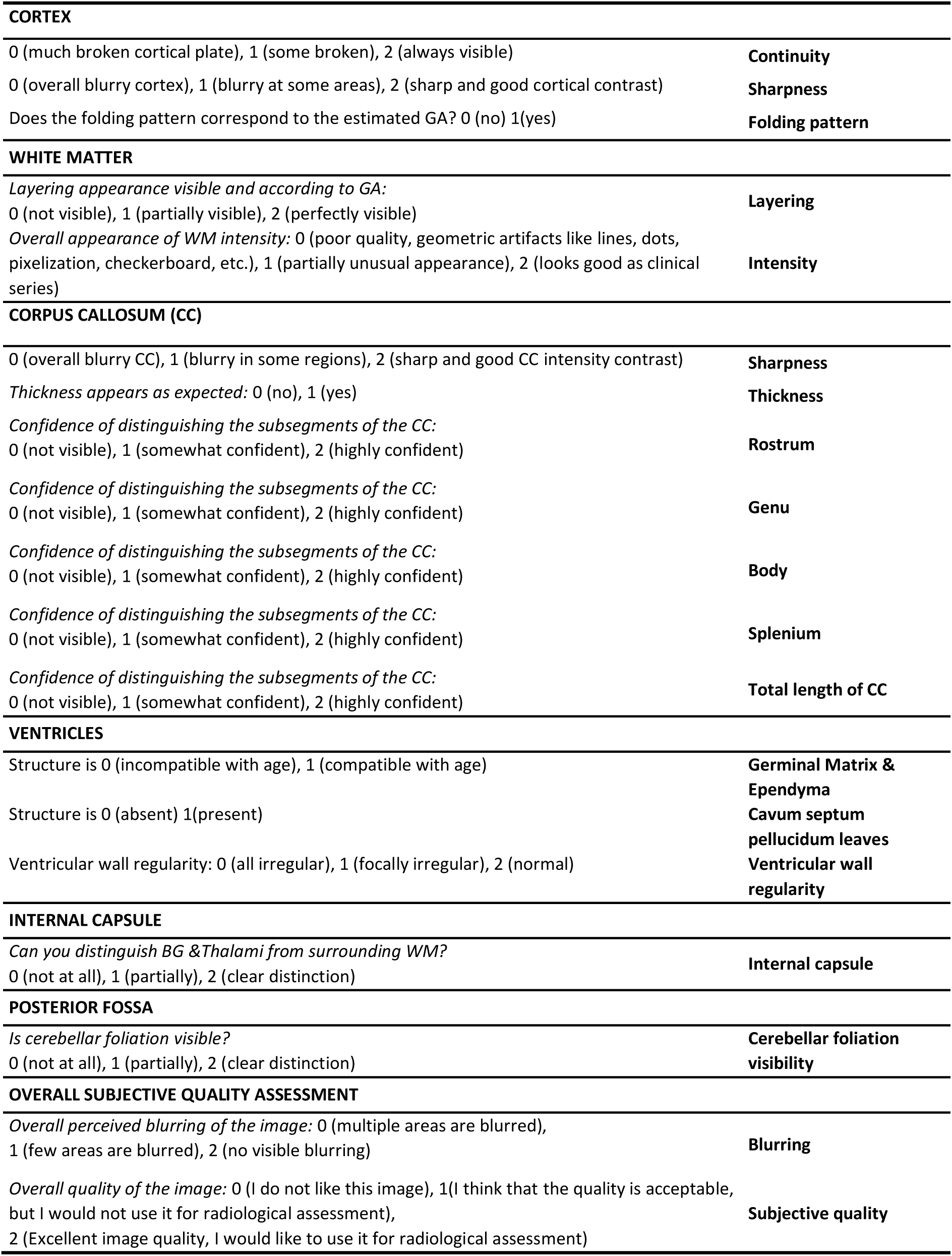
Details of the qualitative ratings asked to the raters in the first stage of the subjective evaluation.

##### Corpus callosum subjective rating

For the corpus callosum, all methods led to a good perception of sharpness and thickness. On the substructures (Table S10A), there was a consistent ordering in the rating quality for all methods (rostrum – genu – splenium/body), independently of the reconstruction method used. On the ventricles, internal capsule and posterior fossa (Table S10B), there was also a consistent hierarchy of NeSVoR < NiftyMIC < SVRTK.

**Table S10.** Subjective structural quality assessment, additional results. **(A)** Assessment of the corpus callosum and the clarity of its substructures on the images. **(B)** Assessment of the ventricles (Is the germinal matrix presence compatible with age; are the cavum septum pellucidum leaves present or absence; is the ventricular wall regular), the internal capsule (Are the basal ganglia (BG) and thalami clearly discernable from the white matter) and the posterior fossa (is the cerebellar foliation clear visible).

| Corpus callosum |  |  |  |  |  |  |  |
| --- | --- | --- | --- | --- | --- | --- | --- |
| (A) | Sharpness | Thickness | Rostrum | Genu | Body | Splenium | Total length CC |
| NeSVoR | 1.04±0.69 | 0.92±0.28 | 0.88±0.90 | 1.25±0.85 | 1.46±0.72 | 1.33±0.70 | 1.29±0.69 |
| NiftyMIC | 1.38±0.71 | 0.83±0.38 | 1.04±0.86 | 1.42±0.83 | 1.63±0.58 | 1.67±0.56 | 1.46±0.66 |
| SVRTK | 1.42±0.58 | 0.83±0.38 | 1.08±0.83 | 1.79±0.51 | 1.71±0.46 | 1.67±0.64 | 1.67±0.48 |
|  |  |  |  |  |  | Posterior Fossa |  |
|  |  | Ventricles |  | Internal capsule |  |  |  |
| (B) | Germinal Matrix & Ependyma | Cavum septum pellucidum leaves | Ventricular wall regularity | BG&Thalami visibility |  | Cerebellar foliation visibility |  |
| NeSVoR | 0.88±0.34 | 0.83±0.38 | 1.25±0.79 | 0.79±0.83 |  | 0.88±0.61 |  |
| NiftyMIC | 0.83±0.38 | 0.83±0.38 | 1.33±0.64 | 1.08±0.78 |  | 1.00±0.78 |  |
| SVRTK | 0.83±0.38 | 0.96±0.20 | 1.46±0.66 | 1.13±0.80 |  | 1.21±0.78 |  |

## Abbreviations

LR: Low-resolution
GA: Gestational Age
SRR: Super-resolution reconstruction
US: Ultrasound
T2w: T2-weighted contrast
LCC: Length of the corpus callosum
HV: Vermis Height
bBIP: Brain biparietal diameter
sBIP: Skull biparietal diameter
TCD: Transverse cerebellar diameter

## Notes

### Competing Interest Statement

The authors have declared no competing interest.

### Author Declarations

The study received ethical approval from each center's institutional review board: Lausanne University Hospital: CER-VD 2021-00124; La Timone: Aix-Marseille University No.2022-04-14-003, Hospital Clinic Barcelona: HCB/2022/0533.

### Summary of Updates

- Correction of typos in some author names - Correction of references to SVRTK

